# A simple non-invasive C reactive protein-based score can predict outcome in patients with COVID-19

**DOI:** 10.1101/2022.09.29.22280522

**Authors:** Riccardo Scotto, Amedeo Lanzardo, Antonio Riccardo Buonomo, Biagio Pinchera, Cattaneo Letizia, Alessia Sardanelli, Simona Mercinelli, Giulio Viceconte, Emanuela Zappulo, Riccardo Villari, Maria Foggia, Ivan Gentile, Federico II COVID-team

## Abstract

**Background:** We evaluated the role of CRP and other laboratory parameters in predicting the worsening of clinical conditions during hospitalization, ICU admission and fatal outcome among patients with COVID-19.

**Methods:** We enrolled consecutive adult inpatients with SARS-CoV-2 infection and respiratory symptoms treated in three different COVID centres. We looked for laboratory parameters collected within 48 hours from hospital admission as predictors of clinical condition.

**Results:** Three-hundred ninety patients were included in the study. At the correlation and regression analysis, age, baseline CRP and LDH were associated with a P/F ratio<200 during hospitalization. At the multivariate analysis, male gender and CRP > 60 mg/l at admission showed to be independently associated with ICU admission. Lymphocytes<1000 cell/μL at admission were associated with worst P/F ratio. The only laboratory predictor of fatal outcome was CRP>60 mg/l at admission. Based on these results, we devised an 11-points numeric ordinary score based on age, sex, CRP and LDH at admission (ASCL score). Patients with ASCL score of 0 or 2 showed to be protected against a P/F ratio<200, while patients with ASCL score of 6, 7 and 8 showed to be at risk for P/F ratio<200. Patients with ASCL score≥7 had a significant increase to die during the hospitalization.

**Conclusions:** Patients with CRP>60 mg/l or LDH>300 IU/l at hospital admission, as well as patients with an ASCL score>6 at hospital admission, should be prioritized for careful respiratory function monitoring and early treatment to prevent a progression of the disease.

## INTRODUCTION

The COVID-19 pandemic is a serious threat for global health, with an unprecedented impact over the last 100 years. On the 8th of June 2022, there were 530,896,347 confirmed cases of COVID-19, including 6,301,020 deaths reported to the WHO (1). After 2 years of pandemic, the efforts in the research headed to the development of vaccines with excellent effectiveness and safety profile in the prevention of COVID-19 (2-4). The availability of both mRNA-based and viral vector vaccines significantly succeeded in minimizing the burden of the pandemic worldwide in terms of severe disease progression, hospitalizations, and deaths (5, 6). Nevertheless, there are sub-groups of patients who remain at high risk for severe disease, intensive care unit (ICU) and death, regardless to vaccination status. (7-10). It is indeed well known that older age, obesity, chronic comorbidities (e.g., diabetes mellitus, cardiovascular and pulmonary diseases, chronic liver and kidney diseases) and immunodeficiency can put patients at high risk for severe COVID-19 (11). These patients can easily progress to the hyperinflammatory phase of SARS-CoV-2 infection (12). At this stage, markers of systemic inflammation are elevated, and SARS-CoV-2 infection can result in a decrease in helper, suppressor, and regulatory T cell counts (13). Studies showed that inflammatory cytokines and biomarkers such as IL-2, IL-6, IL-7, granulocyte colony-stimulating factor, macrophage inflammatory protein 1-a, tumor necrosis factor-a, CRP, ferritin and d-dimer are significantly elevated in those patients with more severe disease (14, 15). (12). A meta-analysis showed that elevated CRP, elevated LDH and lymphopenia were among the most prevalent laboratory findings in patients with COVID-19 (16). In the same study, a correlation between these laboratory abnormalities and the severity of COVID-19 was found. Namely, patients with increased CRP, increased LDH or lymphopenia, were found to be at high risk for severe COVID-19. However, a direct correlation between CRP and LDH levels in COVID-19 patients and the worsening of the respiratory function has not been proven yet. Given such considerations, the aim of the present study was to analyse the correlation between laboratory parameters at admission (including CRP) in patients hospitalized for COVID-19 and the rate of deterioration of the respiratory function after admission.

## MATERIAL AND METHODS

### Study Design

A multicentre retrospective study was conducted among adult inpatients with COVID-19 hospitalized between January 2020 and April 2021 and referring to the following clinical centres:

- Unit of Infectious Diseases. University Hospital Federico II, Naples.
- Hospital “D. Cotugno”. AORN “Dei Colli”, Naples.
- Hospital “G. Rummo”, Benevento.
- Hospital “Sant’Anna e San Sebastiano”, Caserta;

All included patients had a diagnosis of SARS-CoV-2 infection performed with a molecular (PCR) nasal and oropharyngeal swab and were hospitalized for COVID-19-related symptoms. The following exclusion criteria were applied:

- Absence of respiratory symptoms related to COVID-19.
- No serum CRP performed at admission (within 48 hours).
- No serum LDH performed at admission (within 48 hours).
- No arterial blood gas (ABG) test performed at admission (within 48 hours).
- History of a previous SARS-CoV-2 infection or presence of positive SARS-CoV-2 molecular test antecedent 2 weeks from hospitalization.
- History of SARS-CoV-2 vaccination.
- Other hospitalizations in the previous 30 days.

Respiratory symptoms related to COVID-19 included: cough, dyspnoea, tachypnoea, and respiratory failure. Extra-pulmonary manifestations of COVID-19 were not considered for the inclusion in the present study.

The primary outcome of the study was to analyse the correlation between serum CRP at hospital admission and the worst partial pressure of arterial oxygen to fraction of inspired oxygen ratio (P/F ratio) observed during the hospitalization in patients with COVID-19-related respiratory symptoms. Secondary outcomes were:

- To analyse the correlation between serum LDH at hospital admission and the worst P/F ratio observed during the hospitalization.
- To analyse the correlation between blood lymphocyte count at admission and the worst P/F ratio observed during the hospitalization.
- To analyse the presence of risk factors for a worst P/F ratio < 200 during the hospitalization.
- To elaborate a score for prediction of respiratory function deterioration
- To investigate the presence of risk factors for intensive care need during the hospitalization.
- To investigate the presence of risk factors for death during hospitalization.

The clinical records of all included patients were revised, and the following data were collected and reported on an electronic dataset: demographic and clinical data, main comorbidities, laboratory parameters (including CRP, LDH, white blood count), ABGs, outcomes (ICU needs and death). All laboratory parameters were collected at admission (within 48 hours) and every 7 days from admission. All results from ABGs performed during hospitalization were collected and the P/F ratios were calculated. The lowest value of P/F ratio observed during the hospitalization for each patient were collected and reported as “worst P/F ratio”.

The study was conducted according to the guidelines of the Declaration of Helsinki, and approved by the Ethics Committee of University of Naples Federico II (Protocol number 98/2022; Sperimentation I.D. 1032)

### Statistical Analysis

All the variables were tested for parametric/non-parametric distribution with the Kolmogorov-Smirnov test. Comparisons between categorical dichotomous variables were performed with the *χ*^2^ test (or with Fischer’s exact test when applicable), while comparisons between ordinary variables were conducted with the T-student test (parametric variables) or the Mann-Whitney’s U test (non-parametric variables). Comparisons of demographic and laboratory parameters were stratified according to three different clinical outcomes: rate of patients with worst P/F ratio < 200 (meant as the lowest P/F ratio observed during the entire hospitalization for each patient), ICU admission during the hospitalization, and death. The Spearman’s test and the linear regression analysis were used to correlate demographic (age) and laboratory parameters (CRP, LDH, lymphocyte count) with ordinary clinical parameters (namely, worst P/F ratio during hospitalization). The multivariate linear regression analysis was performed including all the parameters significantly correlated with the dependent variable at the univariate linear regression analysis with a p<0.2. In order to identify independent predictors for the three clinical outcomes (worst P/F ratio <200 during hospitalization, ICU admission, death), a logistic regression model was used. Parameters associated with the dependent variables (p<0.2) at the univariate analysis were then included in a multivariate model. A predictive score for worst P/F ratio < 200 during hospitalization was elaborated according to the results of the logistic regression analysis. The age of patients was included in the predictive score based on it was categorized using the same cut-offs of Charlson’s comorbidity index (17). The predictive score was correlated with the worst P/F ratio during hospitalization using the Spearman’s test and logistic regression analysis (ordinary worst P/F ratio) and logistic regression analysis (worst P/F ratio < 200). The diagnostic accuracy for the worst P/F ratio < 200 of the predictive score was evaluated with a ROC curve. For all the tests, a p-value < 0.05 was considered for significance. IBM SPSS© version 27 was used for statistical analysis.

## RESULTS

Globally, 323 patients from the four participant centres were included in the study in accordance with the inclusion/exclusion criteria. Demographic and clinical characteristics of the included patients are reported in Table 1. Most patients showed impaired laboratory parameters within 48 hours from hospital admission, with 35.9% and 50.8% of patients showing CRP values above 60 mg/l and a lymphocyte count below 1000 cell/μl, respectively. Only a minority of patients (4.6%) showed LDH values above 600 IU/l, but 142 patients (44%) had LDH values above 300 IU/l within 48 hours from hospital admission. The median worst P/F ratio observed during the hospitalization was 207 (IQR: 124-301) and nearly half of all the included patients (47.4%) had a worst P/F ratio below 200 during the hospitalization. Rate of ICU admission and death rate were 15.8% and 6.8% respectively. The differences in demographic, clinical and laboratory parameters according to the achievement of the three unfavorable outcomes are shown in Table 2. Patients with a worst P/F ratio below 200 during the hospitalization were more frequently male (p<0.05) and older (p<0.001) than those with P/F ≥ 200. Patients who needed ICU were more frequently male compared to those with no ICU necessity (p<0.001), while those who had a fatal outcome were older than those who survived (p<0.001). Baseline CRP levels were found to be significantly higher among patients with a worst P/F ratio < 200 during the hospitalization (p<0.001) and those who died (p<0.001). Baseline LDH levels were also higher among patients with a worst P/F ratio < 200 during the hospitalization (p<0.001) and those with a fatal outcome (p<0.05). LDH levels were also found to be higher among patients who needed ICU admission (p<0.001). Finally, blood lymphocyte count at admission was lower among patients with a worst P/F ratio during the hospitalization (p<0.001), those who needed ICU (p<0.05) and those who died (p<0.01). Interestingly, the presence of comorbidities was not associated with a worst P/F ratio < 200 nor with ICU admission. However, among patients who survived, most had no comorbidities (p<0.001), while patients who had a fatal outcome, more frequently had 1-2 comorbidities (p<0.05) or 3-5 comorbidities (p<0.01) compared to those who survived. When literature-derived cut-offs for CRP, LDH and lymphocyte blood count were applied, it was found that CRP > 60 mg/l, LDH > 600 IU/l, and lymphocyte < 1000 cell/μl were associated with all the three unfavourable outcomes (worst P/F ratio < 200 during hospitalization, ICU admission, death). Given the paucity of patients with LDH levels above 600 IU/I, a cut-off of 300 IU/l was also applied. The prevalence of patients with LDH > 300 was higher among patients with a worst P/F ratio below 200 during the hospitalization (p<0.001) and those who were admitted to the ICU (p<0.05), compared with patients with a worst P/F ratio above 200 and those who did not needed ICU, respectively. No differences in the rate of patients with LDH > 300 IU/l were found among patients who had a fatal outcome when compared with those who survived.

**Table 1.**
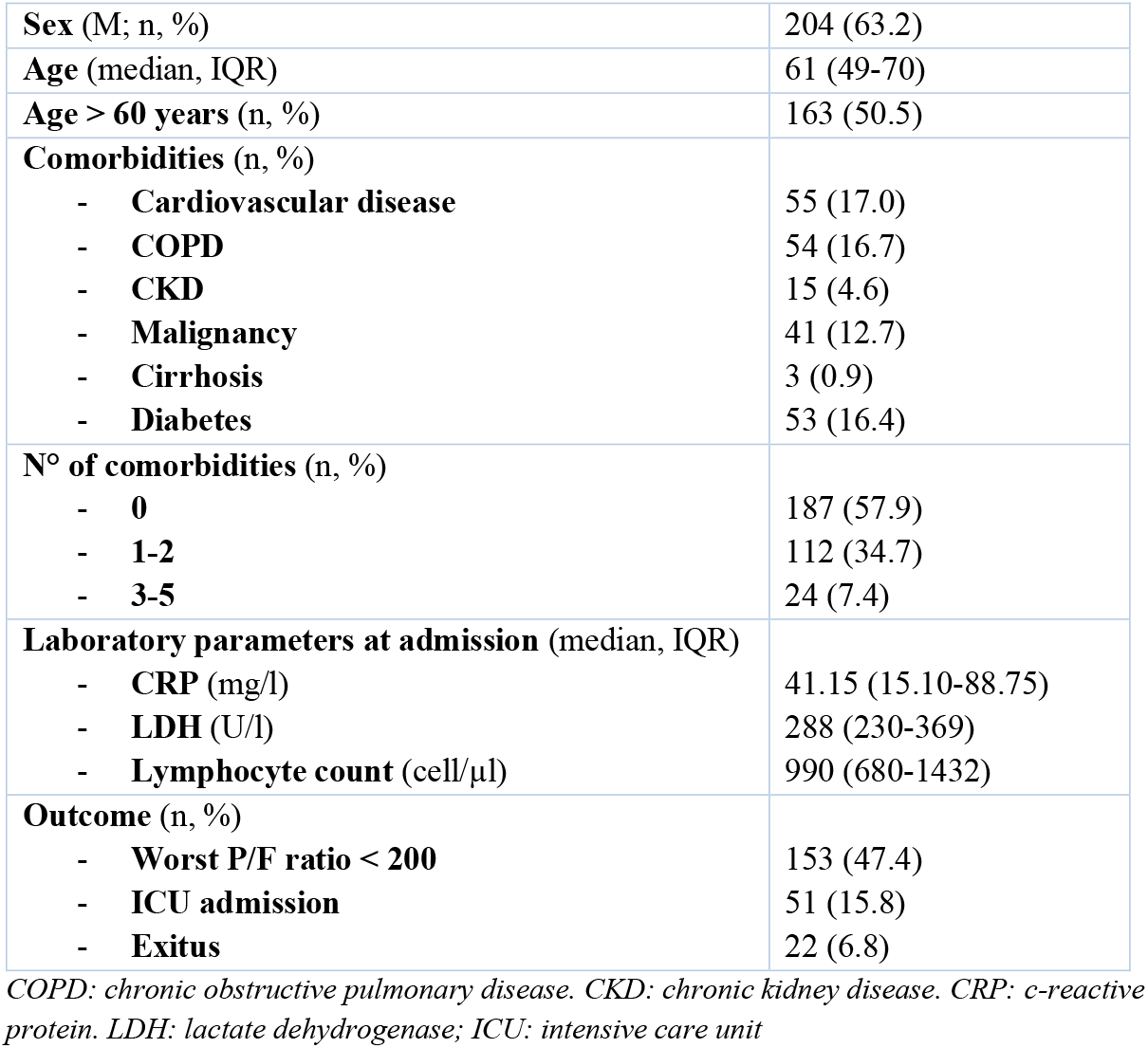
Demographic and clinical characteristics of patients hospitalized for COVID-19 and included in the study (N=323).

|  |  |
| --- | --- |
| **Sex (M; n, %)** | 204 (63.2) |
| **Age (median, IQR)** | 61 (49-70) |
| **Age > 60 years (n, %)** | 163 (50.5) |
| **Comorbidities (n, %)** |  |
| - **Cardiovascular disease** | 55 (17.0) |
| - **COPD** | 54 (16.7) |
| - **CKD** | 15 (4.6) |
| - **Malignancy** | 41 (12.7) |
| - **Cirrhosis** | 3 (0.9) |
| - **Diabetes** | 53 (16.4) |
| **N° of comorbidities (n, %)** |  |
| - **0** | 187 (57.9) |
| - **1-2** | 112 (34.7) |
| - **3-5** | 24 (7.4) |
| **Laboratory parameters at admission (median, IQR)** |  |
| - **CRP (mg/l)** | 41.15 (15.10-88.75) |
| - **LDH (U/l)** | 288 (230-369) |
| - **Lymphocyte count (cell/μl)** | 990 (680-1432) |
| **Outcome (n, %)** |  |
| - **Worst P/F ratio < 200** | 153 (47.4) |
| - **ICU admission** | 51 (15.8) |
| - **Exitus** | 22 (6.8) |

*COPD: chronic obstructive pulmonary disease. CKD: chronic kidney disease. CRP: c-reactive protein. LDH: lactate dehydrogenase; ICU: intensive care unit*

**Table 2:** Differences in the demographic and laboratory parameters among included patients, stratified according to the presence or the absence of three unfavourable outcomes (Worst P/F ratio < 200 during hospitalization, ICU admission, Death)

|  | Worst P/F |  |  | ICU |  |  | Death |  |  |
| --- | --- | --- | --- | --- | --- | --- | --- | --- | --- |
|  | <200 | ≥ 200 | *p-value* | *Yes* | *No* | *p-value* | *Yes* | *No* | *p-value* |
| **Male Sex** (n, %) | 70.9 | 58.1 | **<0.05** | 78.4 | 61.4 | **<0.001** | 68.2 | 63.9 | 0.683 |
| **Age** (median, IQR) | 63 (54-72) | 58 (42-67) | **<0.001** | 65 (52-71) | 60 (49-70) | 0.132 | 78 (71-84) | 60 (48-68) | **<0.001** |
| **Age > 60 years** (n, %) | 56.4 | 37.7 | **<0.001** | 66.0 | 49.2 | **<0.05** | 90.9 | 49.0 | **<0.001** |
| **Comorbidities** (n, %) |  |  |  |  |  |  |  |  |  |
| - **Cardiovascular disease** | 18.3 | 15.9 | 0.564 | 17.6 | 16.9 | 0.898 | 50.0 | 14.6 | **<0.001** |
| - **COPD** | 19.0 | 14.7 | 0.307 | 7.8 | 18.4 | 0.064 | 22.7 | 16.3 | 0.298 |
| - **CKD** | 3.9 | 5.3 | 0.558 | 3.9 | 4.8 | 0.568 | 13.6 | 4.0 | **<0.05** |
| - **Malignancy** | 14.4 | 11.2 | 0.388 | 7.8 | 13.6 | 0.257 | 27.3 | 11.6 | **<0.05** |
| - **Cirrhosis** | 0.7 | 1.2 | 0.625 | 0.0 | 1.1 | 0.596 | 0.0 | 1.0 | 0.638 |
| - **Diabetes** | 17.0 | 15.9 | 0.788 | 15.7 | 16.5 | 0.879 | 45.5 | 14.3 | **<0.001** |
| **N° of comorbidities** (n, %) |  |  |  |  |  |  |  |  |  |
| - **0** | 54.9 | 60.6 | 0.301 | 62.7 | 57.0 | 0.445 | 22.7 | 60.5 | **<0.001** |
| - **1-2** | 37.9 | 31.8 | 0.247 | 35.3 | 34.6 | 0.919 | 54.4 | 33.2 | **<0.05** |
| - **3-5** | 7.2 | 7.6 | 0.876 | 2.0 | 8.5 | 0.081 | 22.7 | 6.3 | **<0.01** |
| **Baseline CRP** (mg/l; median, IQR) | 60.0<br>(21.1-129.9) | 32.0<br>(14.30-60.10) | **<0.001** | 77.4<br>(12.0-137.0) | 39.0<br>(16.0-75.0) | 0.059 | 87.15<br>(45.40-149.0) | 38.5<br>(15.0-80.0) | **<0.001** |
| **Baseline CRP > 60 mg/l** (n, %) | 49.0 | 25.5 | **<0.001** | 52.9 | 33.6 | **<0.01** | 68.2 | 34.4 | **<0.01** |
| **Baseline LDH** (U/l; median, IQR) | 342<br>(256-427) | 269<br>(211-321) | **<0.001** | 357<br>(258-479) | 280<br>(220-351) | **<0.001** | 337<br>(254-479) | 287<br>(228-360) | **<0.05** |
| **Baseline LDH > 600 U/l** (n, %) | 10.1 | 0.0 | **<0.001** | 16.0 | 2.7 | **<0.001** | 13.6 | 4.1 | **<0.05** |
| **Baseline LDH > 300 U/l** (n, %) | 59.7 | 32.3 | **<0.001** | 62.0 | 42.2 | **<0.05** | 59.1 | 44.3 | 0.180 |
| **Baseline Lymphocyte count** (cell/μl; median, IQR) | 861<br>(605-1220) | 1100<br>(720-1550) | **<0.001** | 880<br>(520-1150) | 1000<br>(690-1450) | **<0.05** | 670<br>(430-920) | 1000<br>(690-1440) | **<0.01** |
| **Baseline Lymphocyte count < 1000 cell/μl** (n, %) | 62.2 | 43.4 | **<0.001** | 64.7 | 49.8 | 0.051 | 76.2 | 50.5 | **<0.05** |

ICU: intensive care unit. IQR: interquartile range COPD: chronic obstructive pulmonary disease. CKD: chronic kidney disease. CRP: c-reactive protein. LDH: lactate dehydrogenase

At the correlation analysis, a significant and inverse correlation was found between the worst P/F ratio during hospitalization and: age (Spearman’s r= -0.299, p<0.001); basal CRP values (Spearman’s r= -0.293, p<0.001); basal LDH values (Spearman’s r= -0.363, p<0.001). On the other hand, a direct correlation was found between the worst P/F ratio during hospitalization and basal lymphocyte count (Spearman’s r=0.250, p<0.001). At the linear regression analysis, a significant and negative association was found between worst P/F ratio during hospitalization (dependent variable) and: age (B= -2.372, r^2^=0.125, p<0.001); baseline CRP (B=-0.504, r^2^=0.084, p<0.001, Figure 1); baseline LDH (B=-0.256, r^2^=0.116, p<0.001, Figure 2). The lymphocyte count at admission was not significantly associated with the worst P/F ratio during the hospitalization at the regression analysis. Interestingly, age, baseline CRP values, and baseline LDH values were significantly associated with the worst P/F ratio during the hospitalization at the multivariate linear regression analysis (all p<0.001) (Table 3).

**Table 3:** Univariate and multivariate linear regression analysis between the worst P/F ratio during hospitalization (dependent), age and laboratory parameters at admission

|  | Univariate Analysis |  |  | Multivariate Analysis |  |  |
| --- | --- | --- | --- | --- | --- | --- |
|  | *B* | 95CI | p-value | *B* | 95CI | p-value |
| **Worst P/F ratio<sup>#</sup>** | - | - | - | - | - | - |
| **Age** | -2.372 | -3.073 to -1.672 | **<0.001** | -2.079 | -2.724 to -1.433 | **<0.001** |
| **CRP** | -0.504 | -0.690 to -0.319 | **<0.001** | -0.323 | -0.497 to -0.149 | **<0.001** |
| **LDH** | -0.256 | -0.335 to -0.177 | **<0.001** | -0.205 | -0.279 to -0.130 | **<0.001** |
| **Lymphocyte** | 0.000 | -0.005 to +0.006 | 0.862 | - | - | - |

<sup>#</sup>Worst P/F ratio was set as dependant variable.
*B*: *B* coefficient. *95CI*: 95% confidence intervals. *CRP*: *c*-reactive protein. *LDH*: lactate dehydrogenase

**Figure 1.**
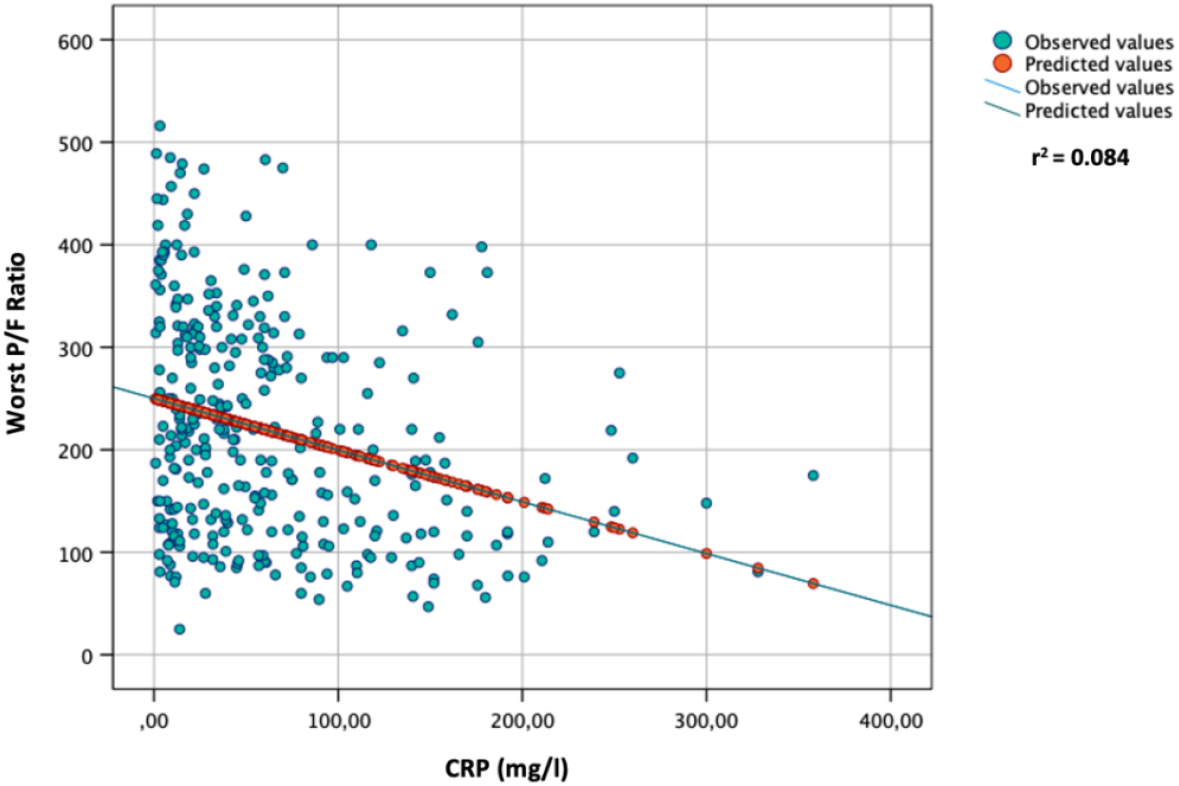
Linear regression analysis between worst P/F ratio during hospitalization (dependent) and CRP levels at admission.

**Figure 2.**
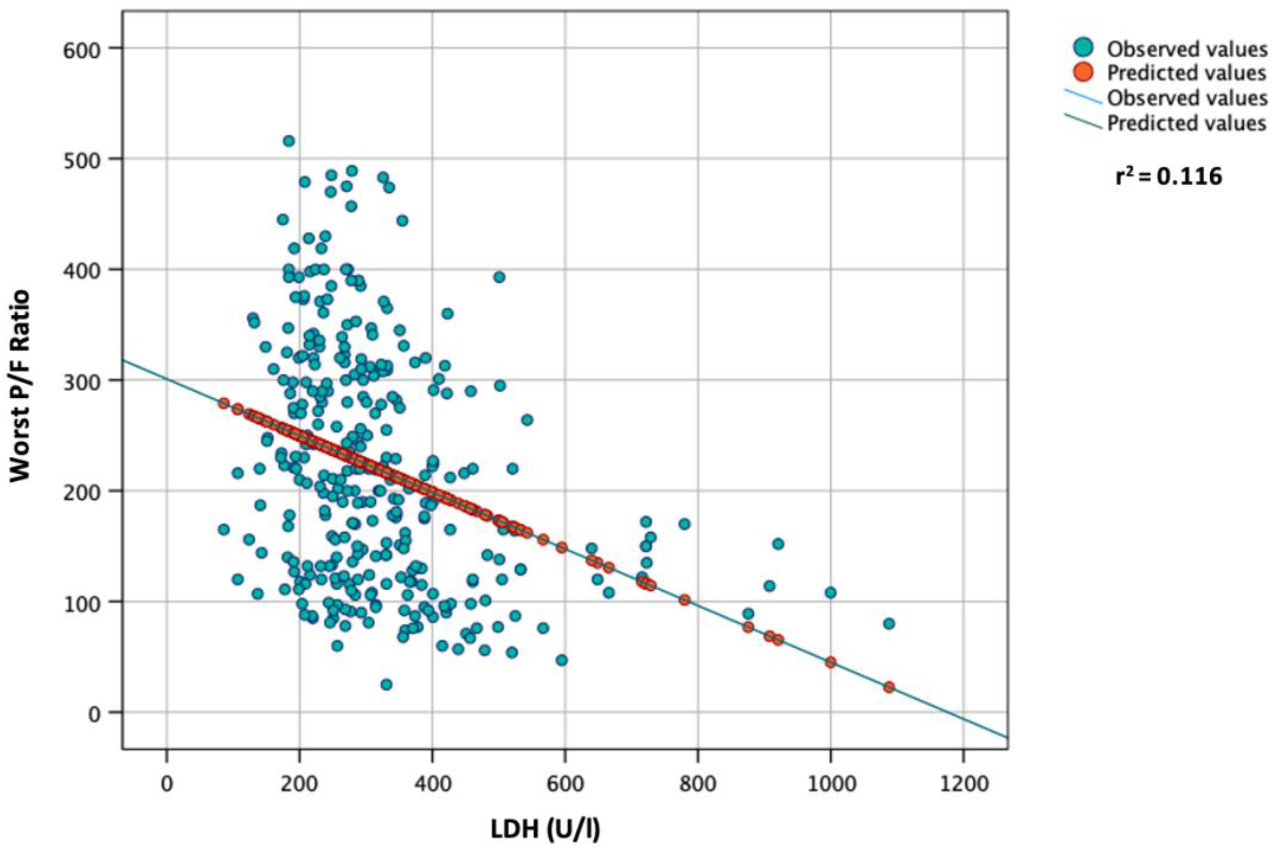
Linear regression analysis between worst P/F ratio during hospitalization (dependent) and LDH levels at admission.

The results of the logistic regression analysis are shown in Table 4. Male sex (aOR 1.73, p<0.05), age > 60 years (aOR: 1.80, p<0.05), CRP > 60 mg/l (aOR: 2.33, p<0.01) and LDH > 300 IU/l (aOR: 2.47, p<0.001) showed to be independently associated with a worst P/F ratio below 200 during the hospitalization. Male sex (aOR: 2.31, p<0.05) and CRP > 60 mg/l at admission (aOR: 2.00, p<0.05) showed to be independently associated with ICU admission at the multivariate analysis. Finally, age > 60 years (aOR: 8.65, p<0.01), the presence of 3-5 chronic comorbidities (aOR: 8.17, p<0.01) and CRP > 60 mg/l at admission (aOR: 5.45, p<0.01) were independently associated with death.

**Table 4:** Univariate and multivariate logistic regression analysis for worst P/F ratio < 200

|  | Univariate Analysis |  |  | Multivariate Analysis |  |  |
| --- | --- | --- | --- | --- | --- | --- |
|  | OR | 95CI | p-value | aOR | 95CI | p-value |
| <u>Worst P/F ratio < 200<sup>#</sup></u> |  |  |  |  |  |  |
| Male sex | 1.75 | 1.10 to 2.80 | **<0.05** | 1.73 | 1.03 to 2.91 | **<0.05** |
| Age > 60 years | 2.14 | 1.36 to 3.56 | **<0.001** | 1.80 | 1.10 to 2.94 | **<0.05** |
| 1-2 comorbidities | 1.31 | 0.83 to 2.08 | 0.247 | - | - | - |
| 3-5 comorbidities | 0.94 | 0.41 to 2.15 | 0.936 | - | - | - |
| CRP > 60 mg/l | 2.81 | 1.75 to 4.52 | **<0.001** | 2.33 | 1.37 to 3.94 | **<0.01** |
| LDH > 300 U/l | 3.11 | 1.95 to 4.93 | **<0.001** | 2.47 | 1.50 to 4.06 | **<0.001** |
| Lymphocyte < 1000 cell/μl | 2.14 | 1.36 to 3.37 | **<0.001** | 1.38 | 0.83 to 2.29 | 0.209 |
| <u>ICU admission<sup>#</sup></u> |  |  |  |  |  |  |
| Male sex | 2.28 | 1.12 to 4.65 | **<0.05** | 2.31 | 1.08 to 4.92 | **<0.05** |
| Age > 60 years | 2.00 | 1.06 to 3.77 | **<0.05** | 1.66 | 0.86 to 3.21 | 0.130 |
| 1-2 comorbidities | 1.03 | 0.55 to 1.93 | 0.919 | - | - | - |
| 3-5 comorbidities | 0.22 | 0.03 to 1.64 | 0.214 | - | - | - |
| CRP > 60 mg/l | 2.22 | 1.21 to 4.08 | **0.01** | 2.00 | 1.03 to 3.86 | **<0.05** |
| LDH > 300 U/l | 2.23 | 1.20 to 4.16 | **<0.05** | 1.74 | 0.89 to 3.41 | 0.107 |
| Lymphocyte < 1000 cell/μl | 1.85 | 0.99 to 3.44 | 0.054 | 1.18 | 0.60 to 2.33 | 0.628 |
| <u>Death<sup>#</sup></u> |  |  |  |  |  |  |
| Male sex | 1.21 | 0.48 to 3.07 | 0.683 | 0.93 | 0.33 to 2.60 | 0.885 |
| Age > 60 years | 10.42 | 2.39 to 45.39 | **<0.01** | 8.65 | 1.86 to 40.33 | **<0.01** |
| 1-2 comorbidities | 2.41 | 1.01 to 5.77 | **<0.05** | 2.85 | 0.92 to 8.87 | **0.07** |
| 3-5 comorbidities | 4.36 | 1.45 to 13.11 | **<0.01** | 8.17 | 1.72 to 38.71 | **<0.01** |
| CRP > 60 mg/l | 4.09 | 1.62 to 10.37 | **<0.01** | 5.45 | 1.82 to 16.34 | **<0.01** |
| LDH > 300 U/l | 1.81 | 0.75 to 4.38 | 0.185 | 1.02 | 0.36 to 2.90 | 0.969 |
| Lymphocyte < 1000 cell/μl | 3.13 | 1.12 to 8.78 | **<0.05** | 2.20 | 0.71 to 6.78 | 0.169 |

<sup>#</sup> Dependant variables.
OR: Odds Ratio. 95CI: 95% confidence intervals. aOR: adjusted Odds Ratio. CRP: c-reactive protein. LDH: lactate dehydrogenase

Given the results from the linear and logistic regression analysis for worst P/F ratio < 200, a 11-points numeric ordinary score based on age, sex, CRP at admission and LDH at admission (ASCL score) was elaborated (Supplementary Table 1). The median ASCL score among patients included in the study was 5 (IQR: 3-6). The highest the ASCL score, the highest was the risk for P/F<200 during the hospitalization (Supplementary Table 2). In particular, patients with ASCL score of 0 (OR: 0.20; 95CI: 0.07 to 0.60) or 2 (OR: 0.43; 95CI: 0.20 to 0.90) showed to be protected against a P/F ratio < 200, while patients with ASCL score of 6 (OR: 2.31; 95CI: 1.18 to 4.52), 7 (OR: 3.30; 95CI: 1.35 to 8.09) and 8 (OR: 2.54; 95CI: 1.16 to 5.59) showed to be at risk for P/F ratio <200. No patients with ASCL score of 1(n=4) had a P/F < 200, while all the patients with ASCL score of 10 (n=5) showed a worst P/F ratio < 200 during the hospitalization. The regression analysis for the worst P/F ratio (dependent variable) showed that for each 1-point increase of the ASCL score, a reduction of the worst P/F ratio of approximately 22 is expected (B=-21.65; 95CI: -26.16 to -17.15, r^2^=0.223, p<0.001) (Figure 3). The diagnostic accuracy of ASCL score for P/F ratio deterioration below 200 was fair (AUC: 0.717, p<0.001) (Supplementary figure 1). Finally, the ASCL score was significantly higher among patients who died (median: 7; IQR: 6-8) compared with patients who survived (median: 4; IQR: 3-6, p<0.001). The diagnostic accuracy of ASCL score for death was good (AUC: 0.804, p<0.001). Patients with ASCL score ≥ 7 had a significantly higher probability to die during the hospitalization (OR: 6.37; 95CI: 2.59 to 15.65, p<0.001) than those with a ASCL less than 7.

**Figure 3.**
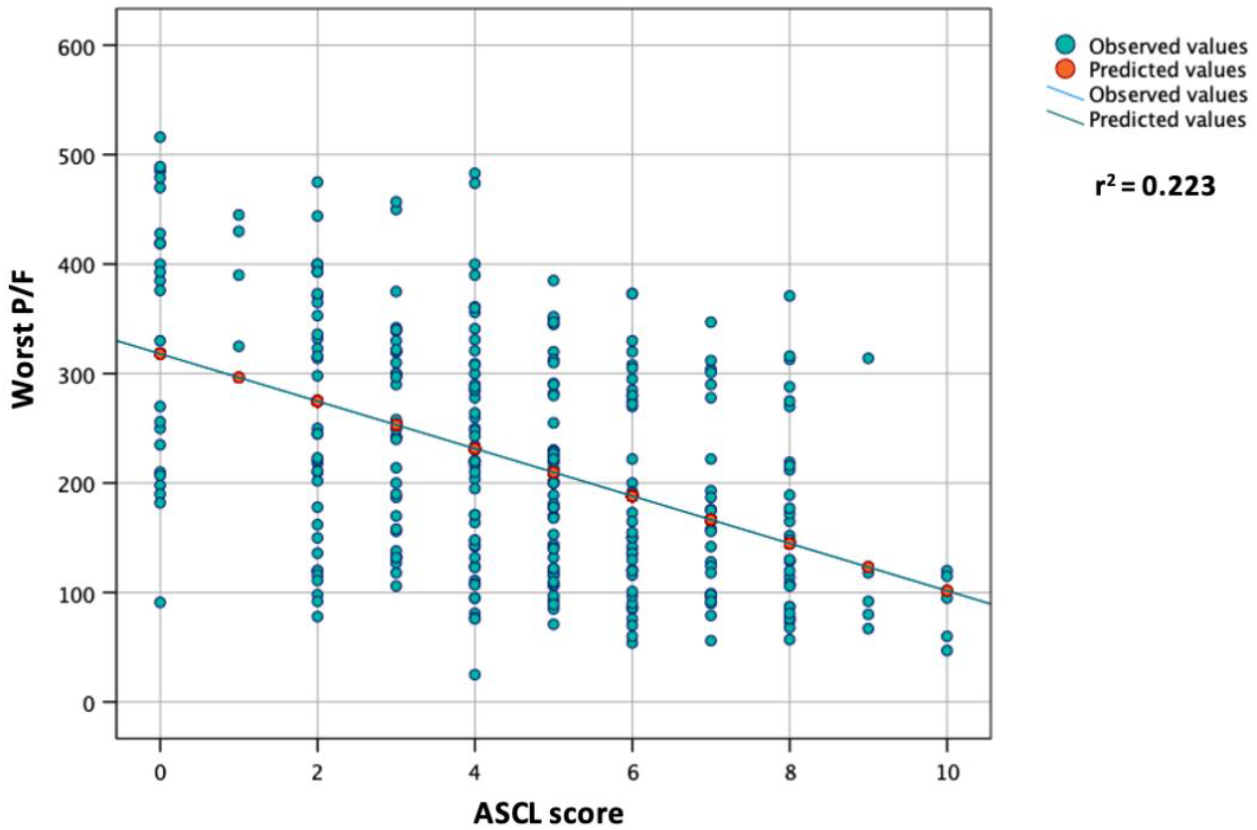
Linear regression analysis between worst P/F ratio during hospitalization (dependent) and ASCL score at admission.

## DISCUSSION

The pandemic of coronavirus disease 2019 (COVID-19) has caused an unprecedented global, social and economic impact, and high numbers of deaths. The clinical features of COVID-19 are diverse and range from asymptomatic to critical illness and death, with severe and critical cases represented by 14% and 5% of laboratory-confirmed COVID-19 patients, respectively (18). A good understanding of the possible risk factors in combination to disease immunopathology associated with COVID-19 severity is helpful for clinicians in identifying patients who are at high risk and require prioritized treatment to prevent disease progression and adverse outcome (19). Risk factors range from demographic factors, such as age (20-22), sex and ethnicity (23, 24), diet and lifestyle habits (25, 26), to underlying diseases (27-32) and complications (33-36). Several laboratory abnormalities were also associated with increased risk of severe COVID-19 and disease progression (37-45). In particular, according to the “Rule-of-6” by *Dickens BSL et al*., the presence with SARS-CoV-2 infection within 48 hours from hospital admission of CRP > 60 mg/l, ferritin > 600 μg/l, and LDH >600 IU/l aided to early identify COVID-19 patients at risk of deterioration to the ICU (46). In our study, we found that patients with respiratory deterioration had higher levels of CRP and LDH, and a lower lymphocyte count compared with patients with a P/F > 200 during the hospitalization. Similar results were obtained comparing laboratory parameters in patients requiring or not ICU admission and in patients who survived compared with those with a fatal outcome, as described in other studies (47-49). Interestingly, in our study, the presence of comorbidities was not associated with a P/F < 200 or with ICU admission. However, cardiovascular disease, CKD, malignancy, and diabetes, as well as the presence of at least 1 comorbidity, were significantly more frequent in patients with a fatal outcome. This result points up that, in contrast to laboratory parameters and other demographic characteristics (e.g., age), the presence of comorbidities does not directly influence respiratory function and mechanics. In fact, we found an inverse and significant association between age (p<0.001) serum CRP (p<0.001, Figure 15) and LDH (p<0.001, Figure 1) and the values of worst P/F ratio during hospitalization at the multivariate analysis (Table 3). Moreover, male sex, age > 60 years, CRP > 60 mg/l, and LDH > 300 IU/l were independently associated with respiratory deterioration (P/F below 200 during the hospitalization). CRP > 60 mg/l was found to be an independent risk factor for ICU admission and death, while LDH > 300 IU/l only showed an association with ICU admission at the univariate logistic regression analysis. The blood lymphocyte count <1,000 cells/μL was not associated with P/F < 200, probably because the white blood count of patients with COVID-19 is considerably influenced by the inflammatory status and, thus, dependant to CRP values. Similarly, no associations were found between lymphocyte < 1,000 cells/μL and ICU admission or death. Finally, the presence of at least 3 comorbidities was found to be an independent risk only for mortality. In accordance with the results from the multivariate logistic regression analysis for a P/F < 200 during hospitalization, we devised a score based on Age, Sex, CRP and LDH (ASCL score, Supplementary Table 1). A progressive increase in the ASCL score was found to be significantly associated with disease progression and respiratory function deterioration, as well as with the risk of death.

There is a spate of literature showing the association between lymphopenia and COVID-19 severity (47, 50). The decreased lymphocyte counts might be caused by viral attachment, immune injuries from inflammatory mediators, or exudation of circulating lymphocytes into inflammatory lung tissues (51).

Elevated serum LDH levels have been widely reported in COVID-19 cases and were predominantly higher in severe patients (52). A meta-analysis showed that the mean value of LDH in severe patients with COVID-19 was 1.54 times higher than in non-severe cases (53). The positive correlation between levels of LDH and disease severity makes it a valuable candidate biomarker for monitoring severe COVID-19 patient. Since higher levels of LDH had been observed in non-survivors at the early stage of illness (48), measuring this parameter at admission will be of greater predictive value for patients’ risk to predict death risk. Elevated LDH values showed to be correlated with the lung injury Murray score in patients with COVID-19 (54) and thus, elevated LDH values at the early stages of SARS-CoV-2 infection can likely predict a severe deterioration of respiratory function.

Finally, elevated CRP is a key marker of disease progression and a risk factor for mortality in COVID-19 patients and it is indicative of developing cytokine storm in COVID-19 patients (20, 55). Out of 32 studies, 20 showed a nearly four-fold higher risk of poor outcomes in COVID-19 patients with elevated CRP (49). Moreover, analysis of patients admitted to the ICU showed an increase of CRP levels in the first seven days (56), suggesting that CRP levels may be correlated with lung injury and respiratory function in patients with COVID-19. Although the role of CRP as predictive factor for disease progression and mortality in patients with SARS-CoV-2 infection has been widely established, a direct correlation between CRP levels and respiratory function is yet to be documented. As an indicator of triggered cytokine storm, elevated CRP levels in the early phases of the infection may predict a subsequent lung damage and respiratory function deterioration caused by the hyper-inflammatory status in patients with COVID-19, as conceivable from the results by *Dickens BSL et al*. (46). Aside from the limited number of patients included in their cohort, the score by *Dickens BSL et al*. did not consider the prognostic weight of demographic factors (such as sex, age, and ethnicity) which are known to heavily influence the prognosis of patients with COVID-19. Finally, the authors did not clarify whether their score was correlated with respiratory function deterioration or other clinical variables.

We acknowledge that this study had some limitations, especially in consideration of its retrospective nature that partially compromised the data collection. In fact, several patients were excluded from the study as they did not perform CRP/LDH nor ABG at admission. Only a minority of patients needed ICU admission (51, 15.8%) or had a fatal outcome (22, 6.8%) and this must be taken into account in interpreting the results. Nevertheless, we believe that the sample size was sufficient to draw significant conclusions regarding correlations with the worst P/F. Moreover, it is known that serum CRP levels in patients with COVID-19 may be affected by the presence of bacterial co-infections. In this cohort of patients, the presence of bacterial co-infections at admission was not systematically evaluated. However, a systematic review and meta-analysis showed that the co-infection rate among patients with COVID-19 is relatively low (7%) (57) and this rate is even lower when considering the presence of bacterial co-infections at hospital admission (3%) (58). Having said that, a routine and systematic screening for bacterial infections at hospital admission in patients with COVID-19 is not recommended, and we believe that the possible presence of bacterial co-infections at hospital admission among patients included in this study cohort unlikely affected the results. Finally, the ASCL score must be validated in more numerous prospective cohorts to draw significant conclusion regarding its diagnostic accuracy in predicting respiratory deterioration in patients with COVID-19.

In conclusion, despite the above-mentioned limitations, the results from this study showed that CRP and LDH levels at admission well correlates with the deterioration of respiratory function in patients with COVID-19. A score based on age, sex, CRP and LDH at admission seems to have a good predictive role in the progression of the respiratory clinical picture and in identifying patients at high risk for unfavourable outcome. Patients with CRP > 60 mg/l or LDH > 300 IU/l at hospital admission, as well as patients with an ASCL score > 6 at hospital admission, should be prioritized for careful respiratory function monitoring and early treatment with specific drugs (i.e., remdesivir, monoclonal antibodies, nirmatrelvir/ritonavir, molnupiravir), when indicated, to prevent a progression of the disease.

## Data Availability

All data produced in the present study are available upon reasonable request to the authors

## Federico II COVID-team

Luigi Ametrano, Francesco Beguinot, Antonio Riccardo Buonomo, Giuseppe Castaldo, Letizia Cattaneo, Maria Carmela Domenica Conte, Mariarosaria Cotugno, Alessia d’Agostino, Giovanni Di Filippo, Isabella Di Filippo, Antonio Di Fusco, Nunzia Esposito, Mariarosaria Faiella, Lidia Festa, Maria Elisabetta Forte, Ludovica Fusco, Antonella Gallicchio, Agnese Giaccone, Anna Iervolino, Carmela Iervolino, Antonio Iuliano, Amedeo Lanzardo, Federica Licciardi, Matteo Lorito, Simona Mercinelli, Fulvio Minervini, Giuseppina Muto, Mariano Nobile, Biagio Pinchera, Giuseppe Portella, Laura Reynaud, Alessia Sardanelli, Marina Sarno, Nicola Schiano Moriello, Maria Michela Scirocco, Fabrizio Scordino, Stefano Mario Susini, Riccardo Scotto, Anastasia Tanzillo, Grazia Tosone, Maria Triassi, Emilia Trucillo, Annapaola Truono, Ilaria Vecchietti, Giulio, Viceconte, Emilia Anna Vozzella, Irene Zotta, Giulia Zumbo.

---

**Supplementary Table 1:**
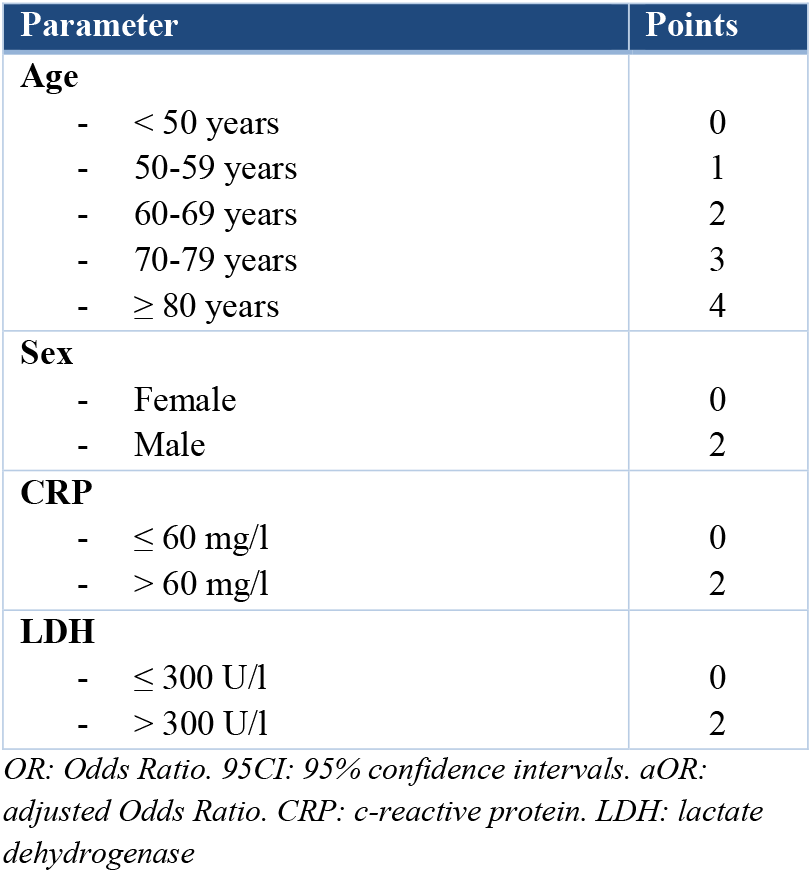
The ASCL score, based on age, sex, CRP at hospital admission and LDH at hospital admission

| Parameter | Points |
| --- | --- |
| **Age** |  |
| - < 50 years | 0 |
| - 50-59 years | 1 |
| - 60-69 years | 2 |
| - 70-79 years | 3 |
| - $\geq 80$ years | 4 |
| **Sex** |  |
| - Female | 0 |
| - Male | 2 |
| **CRP** |  |
| - $\leq 60$ mg/l | 0 |
| - $> 60$ mg/l | 2 |
| **LDH** |  |
| - $\leq 300$ U/l | 0 |
| - $> 300$ U/l | 2 |

OR: Odds Ratio. 95CI: 95% confidence intervals. aOR: adjusted Odds Ratio. CRP: c-reactive protein. LDH: lactate dehydrogenase

**Supplementary Table 2:** Logistic regression analysis for P/F ratio < 200 during the hospitalization according to the ASCL score

| ASCL score | P/F ratio < 200 (n=153) |  |  |
| --- | --- | --- | --- |
|  | %* | OR | 95CI |
| **0** | 16.7 | 0.20 | 0.07 to 0.60 |
| **1** | 0.0 | # | # |
| **2** | 29.7 | 0.43 | 0.20 to 0.90 |
| **3** | 34.3 | 0.54 | 0.26 to 1.13 |
| **4** | 37.7 | 0.62 | 0.34 to 1.14 |
| **5** | 49.0 | 1.08 | 0.59 to 1.97 |
| **6** | 65.1 | 2.31 | 1.18 to 4.52 |
| **7** | 73.1 | 3.30 | 1.35 to 8.09 |
| **8** | 67.7 | 2.54 | 1.16 to 5.59 |
| **9** | 80.0 | 4.54 | 0.50 to 41.04 |
| **10** | 100.0 | ‡ | ‡ |

\*Raw percentage
#Incalculable: no patients with ASCL score = 1 had a P/F ratio < 200
‡Incalculable: all patients with ASCL score = 10 had a P/F ratio < 200
ASCL: age, sex, CRP, LDH. ICU: intensive care unit. OR: odds ratio.

**Supplementary Figure 1.**
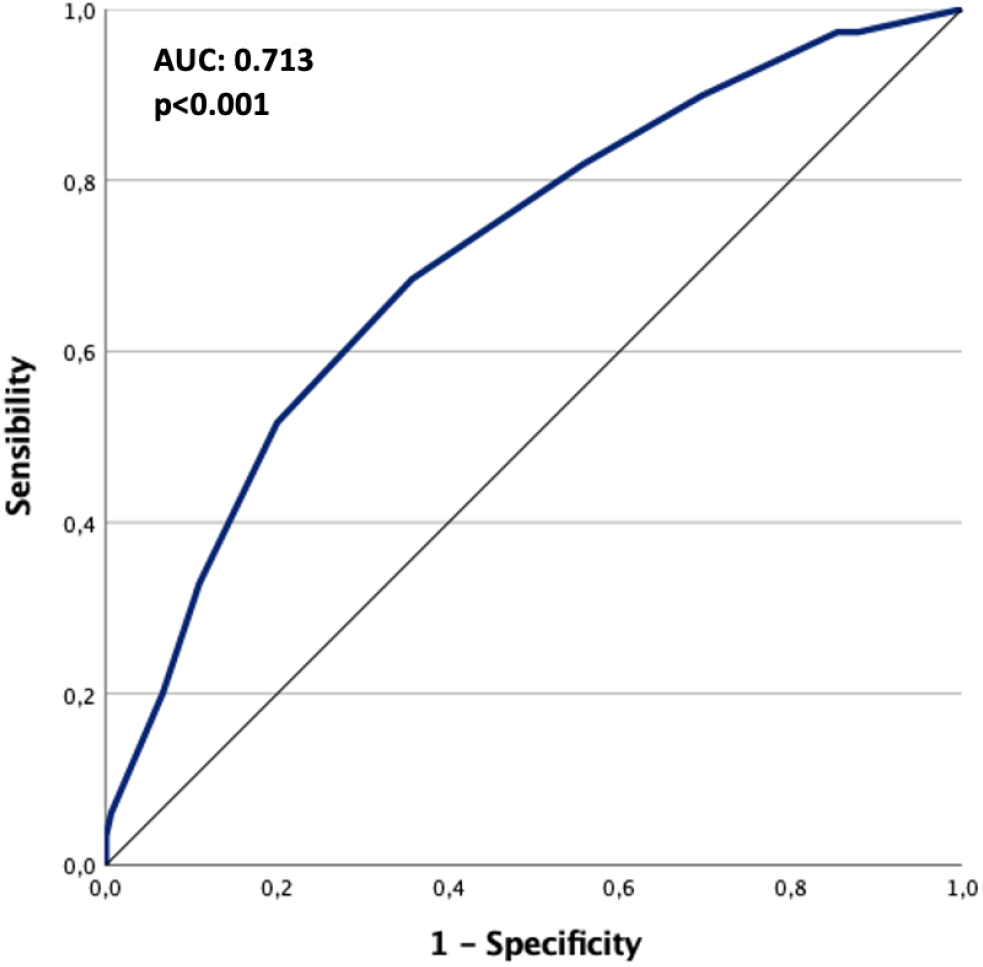
ROC curve for the diagnostic accuracy of the ASCL score in predicting P/F ratio deterioration below 200.

## REFERENCES

1. World Health Organization, WHO Coronavirus (COVID19) Dashboard. Available at: https://covid19.who.int x[Accessed Jun 2021].

2. Polack FP, Thomas SJ, Kitchin N, Absalon J, Gurtman A, Lockhart S, et al. Safety and Efficacy of the BNT162b2 mRNA Covid-19 Vaccine. New England Journal of Medicine. 2020;383(27):2603–15.

3. Baden LR, El Sahly HM, Essink B, Kotloff K, Frey S, Novak R, et al. Efficacy and Safety of the mRNA-1273 SARS-CoV-2 Vaccine. New England Journal of Medicine. 2020;384(5):403–16.

4. Falsey AR, Sobieszczyk ME, Hirsch I, Sproule S, Robb ML, Corey L, et al. Phase 3 Safety and Efficacy of AZD1222 (ChAdOx1 nCoV-19) Covid-19 Vaccine. New England Journal of Medicine. 2021.

5. Moghadas SM, Vilches TN, Zhang K, Wells CR, Shoukat A, Singer BH, et al. The impact of vaccination on COVID-19 outbreaks in the United States. medRxiv : the preprint server for health sciences. 2021:2020.11.27.20240051.

6. Haas EJ, Angulo FJ, McLaughlin JM, Anis E, Singer SR, Khan F, et al. Impact and effectiveness of mRNA BNT162b2 vaccine against SARS-CoV-2 infections and COVID-19 cases, hospitalisations, and deaths following a nationwide vaccination campaign in Israel: an observational study using national surveillance data. Lancet (London, England). 2021;397(10287):1819–29.

7. Herishanu Y, Avivi I, Aharon A, Shefer G, Levi S, Bronstein Y, et al. Efficacy of the BNT162b2 mRNA COVID-19 vaccine in patients with chronic lymphocytic leukemia. Blood. 2021;137(23):3165–73.

8. Collier DA, Ferreira IATM, Kotagiri P, Datir RP, Lim EY, Touizer E, et al. Age-related immune response heterogeneity to SARS-CoV-2 vaccine BNT162b2. Nature. 2021;596(7872):417–22.

9. Yelin I, Katz R, Herzel E, Berman-Zilberstein T, Ben-Tov A, Kuint J, et al. Associations of the BNT162b2 COVID-19 vaccine effectiveness with patient age and comorbidities. medRxiv. 2021:2021.03.16.21253686.

10. Tamuzi JL, Muyaya LM, Mitra A, Nyasulu PS. Systematic review and meta-analysis of COVID-19 vaccines safety, tolerability, and efficacy among HIV-infected patients. medRxiv. 2022:2022.01.11.22269049.

11. Gao Y-d, Ding M, Dong X, Zhang J-j, Kursat Azkur A, Azkur D, et al. Risk factors for severe and critically ill COVID-19 patients: A review. Allergy. 2021;76(2):428–55.

12. Siddiqi HK, Mehra MR. COVID-19 illness in native and immunosuppressed states: A clinical–therapeutic staging proposal. The Journal of Heart and Lung Transplantation. 2020;39(5):405–7.

13. Qin C, Zhou L, Hu Z, Zhang S, Yang S, Tao Y, et al. Dysregulation of Immune Response in Patients With Coronavirus 2019 (COVID-19) in Wuhan, China. Clinical infectious diseases : an official publication of the Infectious Diseases Society of America. 2020;71(15):762–8.

14. Wu C, Chen X, Cai Y, Xia Ja, Zhou X, Xu S, et al. Risk Factors Associated With Acute Respiratory Distress Syndrome and Death in Patients With Coronavirus Disease 2019 Pneumonia in Wuhan, China. JAMA Internal Medicine. 2020;180(7):934–43.

15. Cacciapuoti S, De Rosa A, Gelzo M, Megna M, Raia M, Pinchera B, et al. Immunocytometric analysis of COVID patients: A contribution to personalized therapy? Life Sci. 2020;261:118355.

16. Zhang ZL, Hou YL, Li DT, Li FZ. Laboratory findings of COVID-19: a systematic review and meta-analysis. Scand J Clin Lab Invest. 2020;80(6):441–7.

17. Quan H, Li B, Couris CM, Fushimi K, Graham P, Hider P, et al. Updating and validating the Charlson comorbidity index and score for risk adjustment in hospital discharge abstracts using data from 6 countries. American journal of epidemiology. 2011;173(6):676–82.

18. Wu Z, McGoogan JM. Characteristics of and Important Lessons From the Coronavirus Disease 2019 (COVID-19) Outbreak in China: Summary of a Report of 72 314 Cases From the Chinese Center for Disease Control and Prevention. Jama. 2020;323(13):1239–42.

19. Sokolowska M, Lukasik ZM, Agache I, Akdis CA, Akdis D, Akdis M, et al. Immunology of COVID-19: Mechanisms, clinical outcome, diagnostics, and perspectives-A report of the European Academy of Allergy and Clinical Immunology (EAACI). Allergy. 2020;75(10):2445–76.

20. Zhang JJ, Cao YY, Tan G, Dong X, Wang BC, Lin J, et al. Clinical, radiological, and laboratory characteristics and risk factors for severity and mortality of 289 hospitalized COVID-19 patients. Allergy. 2021;76(2):533–50.

21. Wolff D, Nee S, Hickey NS, Marschollek M. Risk factors for Covid-19 severity and fatality: a structured literature review. Infection. 2021;49(1):15–28.

22. Wang D, Hu B, Hu C, Zhu F, Liu X, Zhang J, et al. Clinical Characteristics of 138 Hospitalized Patients With 2019 Novel Coronavirus–Infected Pneumonia in Wuhan, China. Jama. 2020;323(11):1061–9.

23. Zhang J, Wang X, Jia X, Li J, Hu K, Chen G, et al. Risk factors for disease severity, unimprovement, and mortality in COVID-19 patients in Wuhan, China. Clinical microbiology and infection : the official publication of the European Society of Clinical Microbiology and Infectious Diseases. 2020;26(6):767–72.

24. Ebinger JE, Achamallah N, Ji H, Claggett BL, Sun N, Botting P, et al. Pre-existing traits associated with Covid-19 illness severity. PloS one. 2020;15(7):e0236240.

25. Bousquet J, Anto JM, Czarlewski W, Haahtela T, Fonseca SC, Iaccarino G, et al. Cabbage and fermented vegetables: From death rate heterogeneity in countries to candidates for mitigation strategies of severe COVID-19. Allergy. 2021;76(3):735–50.

26. Bousquet J, Anto JM, Iaccarino G, Czarlewski W, Haahtela T, Anto A, et al. Is diet partly responsible for differences in COVID-19 death rates between and within countries? Clin Transl Allergy. 2020;10:16.

27. Li R, Tian J, Yang F, Lv L, Yu J, Sun G, et al. Clinical characteristics of 225 patients with COVID-19 in a tertiary Hospital near Wuhan, China. Journal of clinical virology : the official publication of the Pan American Society for Clinical Virology. 2020;127:104363.

28. Lighter J, Phillips M, Hochman S, Sterling S, Johnson D, Francois F, et al. Obesity in Patients Younger Than 60 Years Is a Risk Factor for COVID-19 Hospital Admission. Clinical infectious diseases : an official publication of the Infectious Diseases Society of America. 2020;71(15):896–7.

29. Attaway AA, Zein J, Hatipoğlu US. SARS-CoV-2 infection in the COPD population is associated with increased healthcare utilization: An analysis of Cleveland clinic’s COVID-19 registry. EClinicalMedicine. 2020;26:100515.

30. Esposito AJ, Menon AA, Ghosh AJ, Putman RK, Fredenburgh LE, El-Chemaly SY, et al. Increased Odds of Death for Patients with Interstitial Lung Disease and COVID-19: A Case-Control Study. American journal of respiratory and critical care medicine. 2020;202(12):1710–3.

31. Singh S, Khan A. Clinical Characteristics and Outcomes of Coronavirus Disease 2019 Among Patients With Preexisting Liver Disease in the United States: A Multicenter Research Network Study. Gastroenterology. 2020;159(2):768-71.e3.

32. Ng JH, Hirsch JS, Wanchoo R, Sachdeva M, Sakhiya V, Hong S, et al. Outcomes of patients with end-stage kidney disease hospitalized with COVID-19. Kidney International. 2020;98(6):1530–9.

33. Hirsch JS, Ng JH, Ross DW, Sharma P, Shah HH, Barnett RL, et al. Acute kidney injury in patients hospitalized with COVID-19. Kidney Int. 2020;98(1):209–18.

34. Ackermann M, Verleden SE, Kuehnel M, Haverich A, Welte T, Laenger F, et al. Pulmonary Vascular Endothelialitis, Thrombosis, and Angiogenesis in Covid-19. The New England journal of medicine. 2020;383(2):120–8.

35. Nadkarni GN, Lala A, Bagiella E, Chang HL, Moreno PR, Pujadas E, et al. Anticoagulation, Bleeding, Mortality, and Pathology in Hospitalized Patients With COVID-19. Journal of the American College of Cardiology. 2020;76(16):1815–26.

36. Bompard F, Monnier H, Saab I, Tordjman M, Abdoul H, Fournier L, et al. Pulmonary embolism in patients with COVID-19 pneumonia. The European respiratory journal. 2020;56(1).

37. Liu J, Li S, Liu J, Liang B, Wang X, Wang H, et al. Longitudinal characteristics of lymphocyte responses and cytokine profiles in the peripheral blood of SARS-CoV-2 infected patients. EBioMedicine. 2020;55:102763.

38. Huang G, Kovalic AJ, Graber CJ. Prognostic Value of Leukocytosis and Lymphopenia for Coronavirus Disease Severity. Emerging infectious diseases. 2020;26(8):1839–41.

39. Yu HH, Qin C, Chen M, Wang W, Tian DS. D-dimer level is associated with the severity of COVID-19. Thromb Res. 2020;195:219–25.

40. Lippi G, Plebani M. Procalcitonin in patients with severe coronavirus disease 2019 (COVID-19): A meta-analysis. Clin Chim Acta. 2020;505:190–1.

41. Lippi G, Plebani M, Henry BM. Thrombocytopenia is associated with severe coronavirus disease 2019 (COVID-19) infections: A meta-analysis. Clin Chim Acta. 2020;506:145–8.

42. Lei F, Liu YM, Zhou F, Qin JJ, Zhang P, Zhu L, et al. Longitudinal Association Between Markers of Liver Injury and Mortality in COVID-19 in China. Hepatology (Baltimore, Md). 2020;72(2):389–98.

43. Yang X, Jin Y, Li R, Zhang Z, Sun R, Chen D. Prevalence and impact of acute renal impairment on COVID-19: a systematic review and meta-analysis. Critical care (London, England). 2020;24(1):356.

44. Hadjadj J, Yatim N, Barnabei L, Corneau A, Boussier J, Smith N, et al. Impaired type I interferon activity and inflammatory responses in severe COVID-19 patients. Science. 2020;369(6504):718–24.

45. Scotto R, Pinchera B, Perna F, Atripaldi L, Giaccone A, Sequino D, et al. Serum KL-6 Could Represent a Reliable Indicator of Unfavourable Outcome in Patients with COVID-19 Pneumonia. Int J Environ Res Public Health. 2021;18(4).

46. Dickens BSL, Lim JT, Low JW, Lee CK, Sun Y, Nasir HBM, et al. Simple “Rule-of-6” Predicts Severe Coronavirus Disease 2019 (COVID-19). Clinical Infectious Diseases. 2020;72(10):1861–2.

47. Ye W, Chen G, Li X, Lan X, Ji C, Hou M, et al. Dynamic changes of D-dimer and neutrophil-lymphocyte count ratio as prognostic biomarkers in COVID-19. Respir Res. 2020;21(1):169.

48. Wang D, Yin Y, Hu C, Liu X, Zhang X, Zhou S, et al. Clinical course and outcome of 107 patients infected with the novel coronavirus, SARS-CoV-2, discharged from two hospitals in Wuhan, China. Critical care (London, England). 2020;24(1):188.

49. Malik P, Patel U, Mehta D, Patel N, Kelkar R, Akrmah M, et al. Biomarkers and outcomes of COVID-19 hospitalisations: systematic review and meta-analysis. BMJ evidence-based medicine. 2021;26(3):107–8.

50. Guan W-j, Ni Z-y, Hu Y, Liang W-h, Ou C-q, He J-x, et al. Clinical Characteristics of Coronavirus Disease 2019 in China. New England Journal of Medicine. 2020;382(18):1708–20.

51. Wang F, Nie J, Wang H, Zhao Q, Xiong Y, Deng L, et al. Characteristics of Peripheral Lymphocyte Subset Alteration in COVID-19 Pneumonia. The Journal of infectious diseases. 2020;221(11):1762–9.

52. Mori S, Ai T, Otomo Y. Characteristics, laboratories, and prognosis of severe COVID-19 in the Tokyo metropolitan area: A retrospective case series. PloS one. 2020;15(9):e0239644.

53. Bao J, Li C, Zhang K, Kang H, Chen W, Gu B. Comparative analysis of laboratory indexes of severe and non-severe patients infected with COVID-19. Clin Chim Acta. 2020;509:180–94.

54. Liu Y, Yang Y, Zhang C, Huang F, Wang F, Yuan J, et al. Clinical and biochemical indexes from 2019-nCoV infected patients linked to viral loads and lung injury. Sci China Life Sci. 2020;63(3):364–74.

55. Azkur AK, Akdis M, Azkur D, Sokolowska M, van de Veen W, Brüggen MC, et al. Immune response to SARS-CoV-2 and mechanisms of immunopathological changes in COVID-19. Allergy. 2020;75(7):1564–81.

56. Wendel Garcia PD, Fumeaux T, Guerci P, Heuberger DM, Montomoli J, Roche-Campo F, et al. Prognostic factors associated with mortality risk and disease progression in 639 critically ill patients with COVID-19 in Europe: Initial report of the international RISC-19-ICU prospective observational cohort. EClinicalMedicine. 2020;25:100449.

57. Lansbury L, Lim B, Baskaran V, Lim WS. Co-infections in people with COVID-19: a systematic review and meta-analysis. The Journal of infection. 2020;81(2):266–75.

58. Garcia-Vidal C, Sanjuan G, Moreno-García E, Puerta-Alcalde P, Garcia-Pouton N, Chumbita M, et al. Incidence of co-infections and superinfections in hospitalized patients with COVID-19: a retrospective cohort study. Clinical microbiology and infection : the official publication of the European Society of Clinical Microbiology and Infectious Diseases. 2021;27(1):83–8.

